# Cardiac abnormality detection with a tiny diagonal state space model based on sequential liquid neural processing units

**DOI:** 10.1101/2023.12.15.23299990

**Authors:** Zhaojing Huang, Wing Hang Leung, Jiashuo Cui, Leping Yu, Luis Fernando Herbozo Contreras, Nhan Duy Truong, Armin Nikpour, Omid Kavehei

## Abstract

This manuscript presents and studies the performance of the Diagonal State Space Sequence (S4D) model based on the Closed-form Continuous-time (CfC) network in order to achieve a high-performing cardiac abnormality detection method that is robust, generalizable, and tiny in size. Our S4D-CfC model is evaluated on 12- and 1-lead electrocardiogram (ECG) data from over 20,000 patients. The system exhibits validation results with strong average F1 score and average AUROC value of 0.88 and 98%, respectively. To demonstrate the tiny machine learning (tinyML) of our 242 KB size model, we deployed the system on relatively resource-constrained hardware to evaluate its training performance on the edge. Such on-device fine-tuning can enhance personalized solutions in this context, allowing the system to learn each patient’s data features. A comparison with a structured 2D Convolutional LSTM (ConvLSTM2D) CfC model (ConvCfC) demonstrates the S4D-CfC model’s superior performance. The size of the proposed model is also significantly small (25 KB) while maintaining reasonable performance on 2.5s data, 75% shorter than the original 10s data, making it suitable for resource-constrained hardware and reducing latency. In summary, the S4D-CfC model represents a groundbreaking advancement in cardiac abnormality detection, offering robustness, generalization, and practicality with the potential for efficient deployment on limited-resource platforms, revolutionizing healthcare technology.

## I. INTRODUCTION

Currently, numerous methodologies have been devised for the surveillance of cardiac activities, with electrocardiography (ECG) distinguishing itself and receiving extensive utilization owing to its non-invasive characteristics and economic viability. In clinical environments, the 12-lead ECG is the benchmark for gauging the heart’s electrical activity. This methodology necessitates the arrangement of 12-lead, incorporating six limb leads and an additional six precordial leads, to record cardiac dynamics across vertical and horizontal dimensions^1^. To assist clinical practitioners in accurately and swiftly determining the specific locations where patients’ symptoms occur, this study proposes an innovative Neural Circuit Policy (NCP) authorization model. The model effectively reduces the negative impact brought about by the long-term dependency on specific tasks during the learning process, exhibiting a marked advantage over traditional Recurrent Neural Networks (RNNs) such as Long Short-Term Memory (LSTM)^2^. Lechner *et al*. ^2^ demonstrated that the NCP model is a compact representation of neural circuit policy developed from inspirations drawn from known neural computations in the biological brain. Each neuron exhibits substantial computational capacity^3^. In contrast to many deep neural network models that are significantly dependent on clean input data, the NCP model demonstrates greater resilience to the short-term disruptions frequently encountered in real-life scenarios. Furthermore, the NCP model simplifies the entire interpretation process^2^.

Furthermore, this study introduces a Diagonal State Space Sequence (S4D) based model evaluated by our team (Huang *et al*. ^4^) on ECG, which is a derivative of the Structured State Space Sequence (S4) model^5^. The streamlined S4D model reduces computational complexity, significantly enhancing computational efficiency and facilitating quicker analysis of real-time data^4^. This study also utilizes Closed-form Continuous-time Neural Models (CfC)^6^, which efficiently simulate neuron and synapse interactions in neural networks, incorporating liquid time-constant networks (LTC)^3^ within Neural Circuit Policies (NCP)^2^. This paper aims to combine the S4D model with the Closed-form Continuous-time (CfC) network of NCP, proposing a novel S4D-CfC model.

Upon comparison with the previous ConvLSTM2D-CfC (ConvCfC)^7^, improvements in performance and on-chip learning capabilities were observed. Based on the validation data, this model demonstrates superior performance in terms of F1 score and AUROC values compared to previous models. The study’s vision is depicted in Figure 1, aiming to utilize the model in both cloud-based and smart wearable device deployments.

**FIG. 1.**
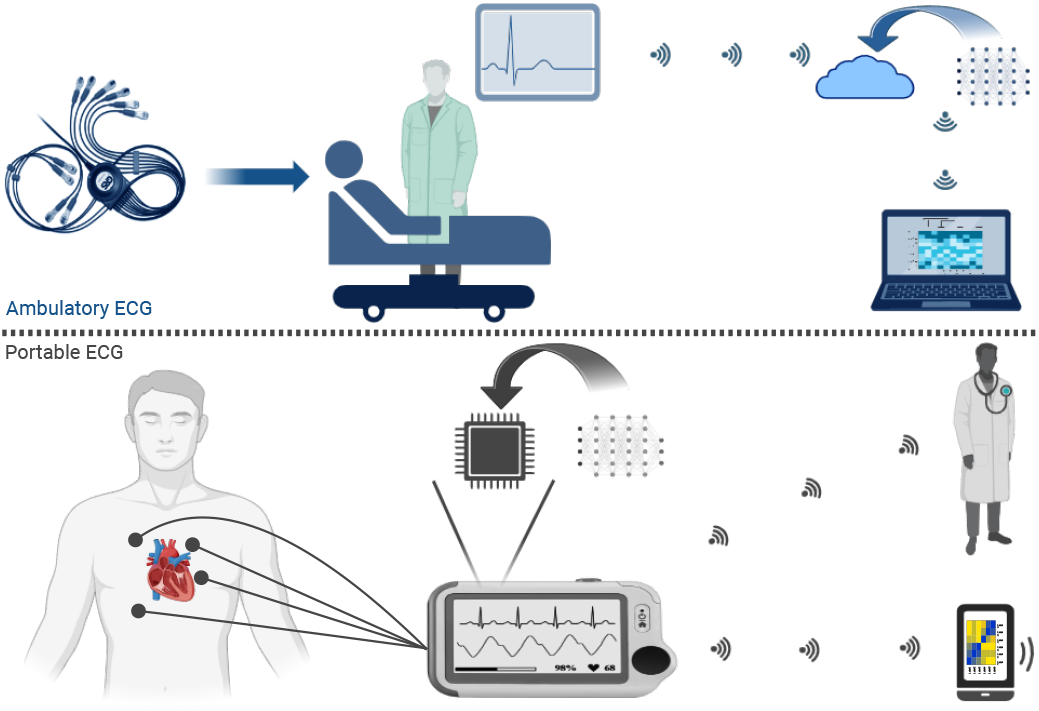
The model’s vision is primarily centered on its utilization in cloud applications and wearable devices. Within this context, its paramount function lies in detecting abnormalities and providing invaluable assistance in their prompt identification.

This study strongly emphasises the model’s compactness for tiny Machine Learning (tinyML) applications. tinyML involves deploying machine learning models on small, resource-constrained devices like microcontrollers and low-power microprocessors^8^. It enables on-device inference (and potentially training), reducing reliance on cloud connectivity. This approach is vital for smart medical sensors, wearables, and computations on edge-of-network applications, extending machine learning capabilities to resource-constrained applications with minimal resources. The model boasts a minimal size of just 242 KB. This compact design not only conserves a significant amount of memory space but also makes efficient use of approximately 70% of the accessible RAM on the Radxa Zero during the on-chip fine-tuning process. Furthermore, it is worth noting that the model can undergo both training and fine-tuning exclusively on the Radxa Zero itself. The model’s performance experiences a notable enhancement post-fine-tuning, underscoring its adaptability and impressive resource efficiency.

### A. Background

The Telehealth Network of Minas Gerais (TNMG) dataset is one of the electrocardiogram datasets extensively used for cardiac rhythm classification^9^. In this context, Zvuloni *et al*. ^10^ compared the performance of Feature Engineering (FE), Deep Learning (DL), and a combined approach (FE+DL) for ECG classification. The FE approach involves initial signal analysis using feature engineering methods to extract features, followed by feature selection using Minimum Redundancy Maximum Relevance (mRMR). In contrast, DL employs deep neural network architectures. FE+DL combines both methods for enhanced ECG classification. Divergently, Biton *et al*. ^11^ integrated features from Heart Rate Variability (HRV), Electrocardiogram Morphology (MOR), Deep Neural Networks (DNN), and patient metadata (META) to enhance the precision and reliability of cardiac disease risk prediction. Differing from the former methodologies, Huang *et al*. ^12^ introduced an attention layer, incorporating ReLU and Softmax mechanisms, to the DNN model framework for abnormality classification by Ribeiro *et al*. ^9^, aiming to study the generalization impact of the attention mechanism on model performance. Meanwhile, Montenegro, Peixoto, and Machado ^13^ improved rhythm classification performance by employing an open-access 1D-CNN model and exploring transfer learning on a smaller database. Huang *et al*. ^7^ introduced the ConvLSTM2D-CfC (ConvCfC) model, which incorporates Convolutions for Cardiac abnormality detection (CfC). Notably, ConvCfC shares similarities with our proposed S4DCfC model, as both models integrate CfC into their architectural design. Consequently, we employ ConvCfC as a baseline for assessing the performance of our proposed model.

Lechner *et al*. ^2^ intended to utilize the NCP model to optimize camera inputs in the context of automotive driving applications. In contrast to conventional Convolutional Neural Networks (CNNs), which often prioritize roadside features while neglecting the inherent characteristics of the road surface, this model presented an innovative approach that placed heightened importance on the road horizon. This significantly enhanced the understanding and decision-making in automotive driving technology. The S4D model, an enhanced version of S4, achieves computational performance comparable to S4 with only two lines of encoded kernels. It has shown advanced results in various fields such as medical time series and audio imaging^4^. The Closed-form Continuous-time (CfC) Neural Networks proposed by Hasani *et al*. ^6^ overcome the limitations of numerical differential equation solvers. Compared to networks based on differential equations, they exhibit a significant improvement in training and inference speed, exceptional scalability, and outstanding performance in time-series modeling tasks. Consequently, the integration of S4D and CfC for developing a new model holds considerable potential in the field of deep learning.

In recent years, the integration of compact neural network algorithms with wearable medical devices has emerged as a significant innovation in the field of medical technology. This technology enables real-time health monitoring, assists physicians in locating abnormalities, and accurately predicts outcomes, thereby substantially improving patient treatment efficacy. Consequently, the on-device training and prediction of compact neural network algorithm models have progressively become a popular research topic^14^.

## II. PREREQUISITE

## A. Diagonal State Space Sequence (S4D) Model

The Structured State Space sequence model (S4) is a dedicated deep learning approach designed to excel in processing extensive data sequences. Its potential becomes evident when dealing with complex datasets marked by intricate patterns and interconnections, as highlighted in^5^. S4 presents itself as a promising and innovative substitute for conventional sequence models such as RNNs, CNNs, or Transformers, as detailed in^5^.

Fundamentally, the continuous-time state-space model operates as a transformation mechanism. The equation is shown in equation 1 and equation 2. It takes a one-dimensional input signal, denoted as *u*(*t*), and transforms it into a higher-dimensional latent state, *x*(*t*), which is subsequently mapped onto a one-dimensional output signal, *y*(*t*). The mathematical representation of this transformation can be expressed through the following equations:

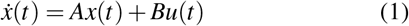

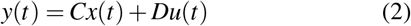

The S4 model is intricately designed to capture the complex temporal dependencies and structural intricacies inherent in time series data. It achieves this through the utilization of a structured state-space model framework. To effectively represent the state matrix and facilitate the modeling of hierarchical dependencies, the model employs a parameterization technique known as the diagonal plus low-rank (DPLR) matrix, which is thoroughly explained in the work of Gu *et al*. ^15^. Conversely, the S4D model takes a streamlined approach to parameterization, as detailed in Gu *et al*. ^15^. This simplified methodology exclusively represents the state matrix as a diagonal matrix, eliminating the low-rank component. This simplification significantly reduces computational complexity, thereby improving the model’s practicality and interpretability. By discarding the low-rank element in the parameterization, the S4D model achieves a more straightforward mathematical representation when compared to its S4 counterpart.

Huang *et al*. ^7^ have successfully built a multiplayer S4D model for ECG data, demonstrating remarkable performance in detecting abnormalities. Building upon their work, we have further developed our proposed model.

### B. Neural circuit policies (NCP)

The Neural Connectivity and Processing (NCP) model represents a comprehensive end-to-end learning framework characterized by its incorporation of multiple convolution layers, as detailed in the research conducted by Lechner *et al*. ^2^. This model is deeply rooted in the intricate neural circuitry of the elegans nematode, an organism that has been extensively studied in the work of Lechner and colleagues^2^.

Within the NCP model, four distinct neural strata serve critical functions: sensory neurons (*N*_*s*_), inter-neurons (*N*_*i*_), command neurons (*N*_*c*_), and motor neurons (*N*_*m*_). A specific number of synapses are introduced to facilitate the transmission of information across these neural layers. A set of mechanisms regulates the establishment of synaptic connections:

- In the context of neural layers, information transmission from source neurons to target neurons occurs via a predefined number of synapses (*n*_*so*−*t*_). These synapses follow a Bernoulli distribution characterized by a probability denoted as *p*_2_^2^.
- When target neurons initially lack synapses, a supplementary set of synapses (*m*_*so*−*t*_) is introduced from source neurons. The quantity of these synapses is determined by a Binomial distribution characterized by a probability denoted as *p*_3_, with the synapses being randomly selected from a pool of source neurons^2^.
- Command neurons exhibit recurrent connections, marked by establishing synapses (*l*_*so*−*t*_) targeting command neurons. These connections are selected by a Binomial distribution with a probability represented as *p*_4_^2^.

Together, these processes oversee information transmission and play a crucial role in shaping the computational dynamics observed within the NCP model, as discussed in^2^.

Equation 3 demonstrates how the semi-implicit Euler method is employed in the context of the NCP model. Within this equation, *I*_*in*_ represents a group of neurons serving as inputs to neuron *i*:

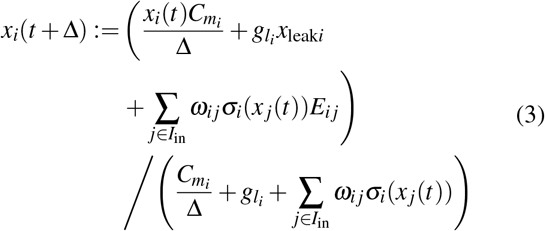

### C. Closed-form Continuous-time (CfC) Neural Networks

The concept of Closed-form Continuous-time (CfC) Neural Networks, as explored in the research conducted by Hasani *et al*. ^6^, is characterized by a unique trait: it operates without the need for numerical solvers in solving Ordinary Differential Equations (ODEs) to generate temporal rollouts. These networks embody sought-after features like flexibility, causality, and continuous-time dynamics, typically associated with ODE-based networks, all while showcasing superior computational efficiency^6^. Equation 4 provides the mathematical representation of the CfC model, where *σ* (−*f* (*x, I*; *θ*_*f*_)*t*) and [1−*σ* (− [*f* (*x, I*; *θ*_*f*_)]*t*)] serve as the mechanisms governing time-continuous gating^6^.

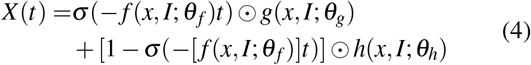

To sum up, the Closed-form Continuous-time (CfC) Neural Networks, as presented by Hasani *et al*. ^6^, present an innovative method for temporal modeling. They do so by avoiding the use of numerical ODE solvers, effectively blending the advantages of ODE-based networks with improved computational efficiency, thanks to their unique time-continuous gating mechanisms.

## III. DATASETS

In our research endeavor, we relied upon two distinct datasets as integral components for evaluating the proposed models. The first dataset, renowned as the CPSC dataset or the 12-lead ECG dataset, underwent meticulous curation for The China Physiological Signal Challenge in 2018, as delineated in^16^. Its primary objective centered around enabling the automated detection of anomalies in both the rhythm and morphology of 12-lead electrocardiograms (ECGs).

In contrast, the second dataset employed in this study originates from the Telehealth Network of Minas Gerais (TNMG) dataset, as expounded in^9^. This dataset predominantly served as the foundational source for training our models.

To comprehensively evaluate the models’ proficiency in handling new and previously unobserved data, we strategically opted for the CPSC dataset as an independent test dataset. This meticulous evaluation was designed to assess the models’ adaptability when confronted with real-world data, markedly distinct from the characteristics of the TNMG dataset.

The central objective of this academic inquiry was to gauge the models’ capacity to generalize and their overall efficacy when applied to unfamiliar data. This evaluation process commenced with the initial training of the models using the TNMG dataset and culminated in a rigorous assessment of their performance on the CPSC dataset. It is of paramount importance to underscore that the outcomes of this evaluation hold profound implications for establishing the models’ reliability and effectiveness in practical, real-life scenarios.

### A. TNMG Dataset

In the realm of our research endeavor, we harnessed the immense potential of the TNMG dataset, a vast repository housing a meticulously curated collection of 2,322,513 labeled samples of 12-lead electrocardiogram (ECG) data. Within this extensive dataset, we encountered six distinct categories of abnormalities: Atrial Fibrillation (AF), Left Bundle Branch Block (LBBB), First Degree Atrioventricular Block (1dAVb), Right Bundle Branch Block (RBBB), Sinus Tachycardia (ST), and Sinus Bradycardia (SB), as detailed in^9^. The original ECG data was recorded at a sampling frequency of 400 Hz.

We executed a systematic approach to construct a well-balanced dataset suitable for model training. Specifically, we conducted random samplings, selecting 3,000 data samples for each of the six aforementioned abnormalities. Additionally, an equivalent number of 3,000 samples exhibiting no indications of abnormalities were incorporated into the dataset. Consequently, this comprehensive data subset consisted of a carefully chosen total of 21,000 instances. In cases where patients presented with multiple abnormalities, any remaining samples needed to reach the predefined subset size of 21,000 were randomly selected from the TNMG dataset.

The dataset underwent a rigorous normalization process, resulting in a standardized length consisting of precisely 4096 readings. This stringent uniformity was meticulously enforced to ensure consistent data analysis and modeling. Any readings exceeding this predetermined length were systematically excluded, streamlining data processing and facilitating meaningful comparisons.

As illustrated in Figure 2(a), a balanced representation of genders is evident within the subset, underscoring the importance of inclusivity and the validation of subsequent analyses. Furthermore, this subset closely mirrors the age distribution observed in the general population, further enhancing its representativeness for age-related investigations.

**FIG. 2.**
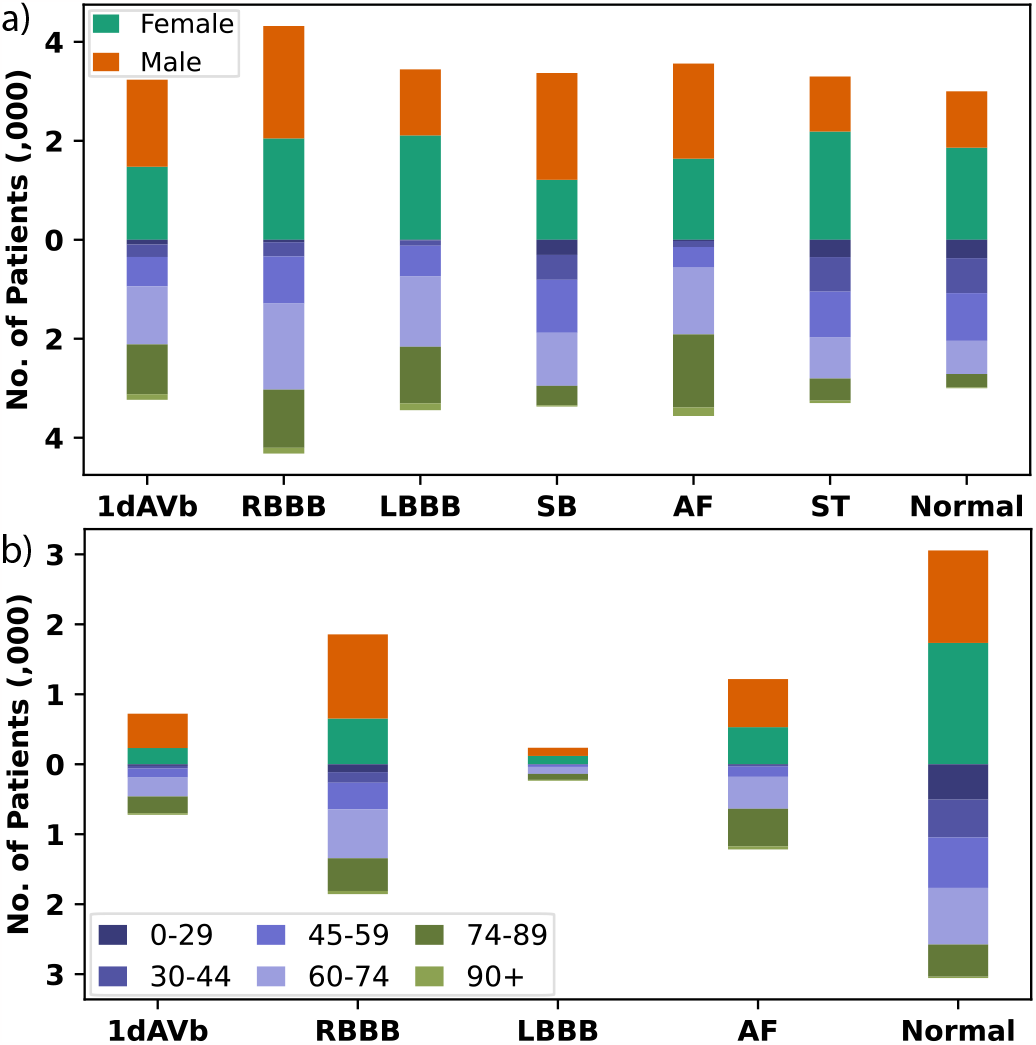
Gender breakdown above, age below in each sub-chart. a) Balanced TNMG subset with six abnormality categories. b) In-depth analysis of the CPSC dataset on studied abnormalities.

Moreover, the sampling strategy employed yielded a well-calibrated distribution of various abnormalities, enabling a comprehensive exploration of their characteristics and impacts. This equilibrium within the dataset significantly enhances the model’s learning process, ultimately contributing to its overall performance improvement, as emphasized in^12^.

### B. CPSC Dataset

The CPSC dataset initially comprises 12-lead electrocardiograms (ECGs) sampled at a rate of 500 Hz. To ensure alignment with the TNMG data, we conducted a resampling operation on this dataset, adjusting it to a sampling rate of 400 Hz to match that of TNMG. This dataset distinguishes itself by incorporating ECGs from patients diagnosed with various cardiovascular conditions, encompassing common cardiac rhythms. Every ECG within this dataset has undergone meticulous expert labeling, resulting in precise annotations for the detected abnormalities. In summary, the dataset encompasses eight distinct categories of abnormalities.

To assess the model’s generalization capabilities, our evaluation focused specifically on four particular abnormalities from this dataset: AF, 1dAVb, RBBB, and LBBB. It’s crucial to emphasize that this study deliberately excluded four other types of abnormalities, namely ST-segment Depression (STD), ST-segment Elevated (STE), Premature Atrial Contraction (PAC), and Premature Ventricular Contraction (PVC). This selection was driven by the fact that only these four abnormalities align with the characteristics found in the TNMG dataset, thereby guiding our choice for evaluation.

In our meticulous data selection process, we applied stringent criteria to eliminate entries in the dataset containing any missing readings. This rigorous curation resulted in a final dataset comprising 6,877 distinct ECG tracings. Subsequently, we standardized the data to a consistent length of 4,096 readings, with any readings exceeding this specified length meticulously removed during the data cleaning phase. For a more comprehensive understanding of the refined dataset, please refer to the detailed information provided in Figure 2(b).

A thorough examination of the dataset revealed a notable gender imbalance, with a higher representation of male patients compared to their female counterparts. However, it’s essential to note that the age distribution of the patients in the dataset closely mirrors that of the general population, with a significant portion belonging to older age groups. Nevertheless, upon closer scrutiny of the distribution of abnormalities, a minor imbalance becomes apparent. Specifically, the prevalence of LBBB is relatively less frequent compared to the occurrence of other abnormalities present in the dataset.

## IV. METHODS

Our central aim is centered on developing an agile and highly efficient model, precisely designed for the analysis of electrocardiogram (ECG) data. Our ultimate aspiration is to optimize this model for efficient hardware utilization and to enable potential deployment in cloud environments, ensuring seamless compatibility with resource-constrained devices. Furthermore, we embark on an exhaustive assessment of the model’s capabilities, thoroughly examining its potential for generalization and its ability to withstand incomplete data scenarios.

In pursuit of this aim, we have conceived a compact architecture that significantly reduces computational demands while upholding precision.

Following this architectural development, we proceed to input the data into the proposed models, primarily for the purposes of training and validation. Subsequently, we subject the model to a rigorous performance evaluation, leveraging carefully curated performance metrics to gain a deeper understanding of its overall effectiveness.

### A. Preprocessing of ECG data

The use of an S4D layer as the input layer for the compact model suggests that pre-processing of ECG data may not be necessary^4^. However, this study will still explore the effect of applying max-min scaling to the data on the model’s performance.

### B. Diagonal State Space Sequence (S4D)

The S4D model, as explained in^15^, utilizes a state matrix representation that solely consists of a diagonal matrix, eliminating the low-rank component. This simplification leads to a notable reduction in computational complexity, rendering the model more suitable for practical implementation and comprehension. Removing the low-rank term in parameterization results in a more straightforward mathematical representation for the S4D model.

### C. Neural Circuit Policies (NCP)

Drawing inspiration from the Caenorhabditis elegans nematode, Neural Circuit Policies (NCPs) have emerged as intelligent agents guided by principles inspired by the human brain. Unlike traditional deep models, NCPs empower individual neurons with advanced computational capabilities. Extensive research highlights that the integration of NCPs results in more streamlined networks, thereby enhancing interpretability in comparison to conventional models, as detailed in^2^.

NCP networks comprise Liquid Time Constant (LTC) neurons^3^. These neurons, designed following NCP principles, result in highly compact, sparsely interconnected networks. LTC neurons, resembling leaky integrators, accumulate input information over time and gradually release stored charge. This mechanism prevents membrane potential saturation, facilitating temporal information processing^2^.

Moreover, in the work of Hasani *et al*. ^6^, they introduced Closed-form Continuous-time Neural Networks (CfC) as a groundbreaking method for time-series modeling. Building upon the concept of liquid networks, CfC models outperform even the most advanced recurrent neural networks by exploiting closed-form ordinary differential equations (ODEs). In accomplishing this, they provide a definitive solution to longstanding integral challenges linked to liquid time-constant dynamics, as comprehensively outlined in^6^.

### D. Simple Model Design

The depicted model architecture (Figure 3) is intentionally crafted to be straightforward yet highly efficient. It primarily comprises a solitary S4D layer, tasked with feature extraction, and feeds into 128 neurons that serve as input for the NCP network. The NCP network is constructed using a CfC arrangement, giving rise to a unique model known as the S4DCfC model. This model encompasses 14 interconnected interneurons and command neurons, along with 6 output neurons. It’s worth noting that the 128 neurons originating from the output of the S4D layer, which then serve as the input neurons for the NCP network, are governed by an internal parameter referred to as the model dimension (d_model). This internal hyper-parameter wields control not only over the number of neurons in the output but also over the count of output channels within the S4D layer. In our comprehensive study, we have discovered that this parameter plays a pivotal role in determining the model’s size, a topic we elaborate on in the results section. The Model Dimension employed throughout the majority of this study is 128.

**FIG. 3.**
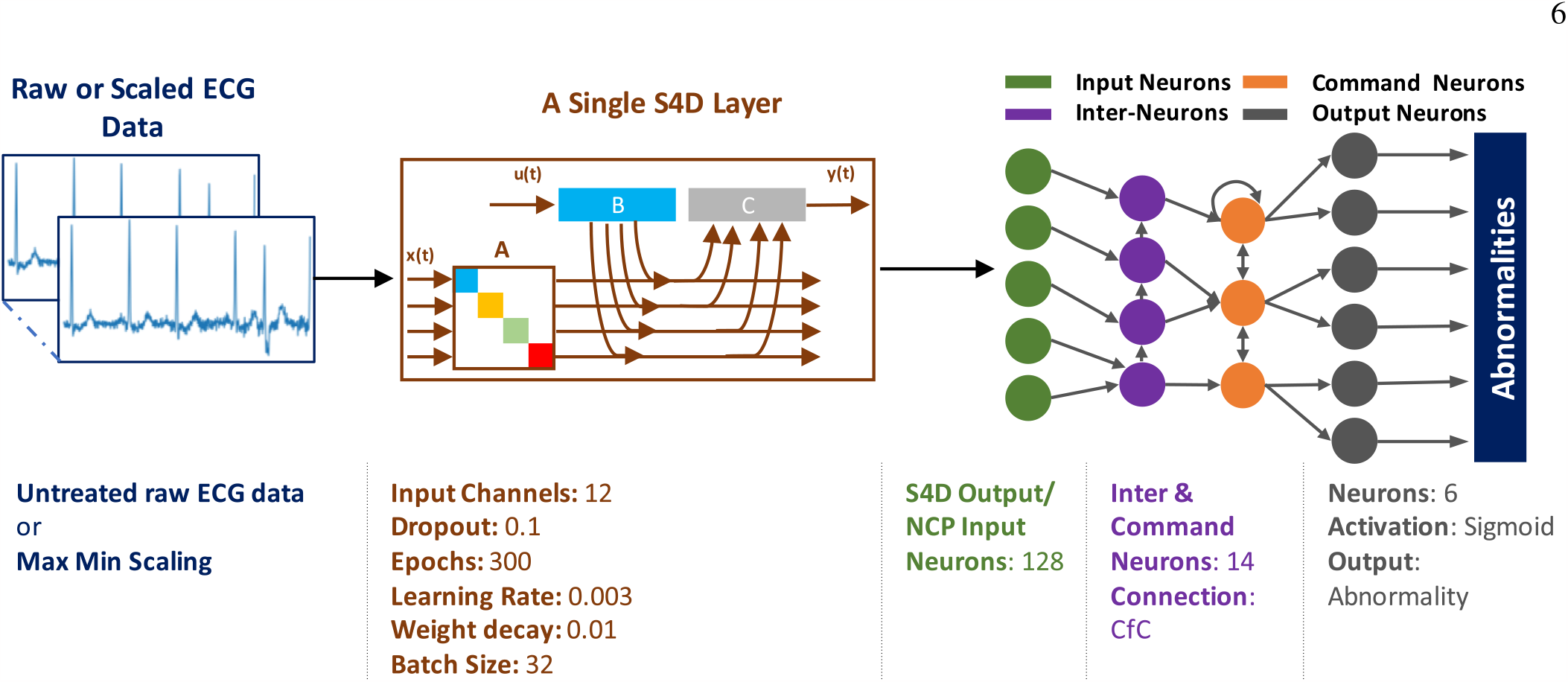
The model architecture is comprised of a core S4D layer designed to process both raw and scaled ECG data. Within this layer, relevant features are extracted and subsequently funneled into a dense layer consisting of 128 neurons, which serve as the input neurons for the NCP network. The connections between input, motor, and output neurons are established through CfC configurations, culminating in the application of a sigmoid activation function at the final stage of the model.

### E. Performance Metrics

Precision and recall serve as critical metrics for evaluating model performance. Precision assesses the accuracy of positive predictions, while recall measures the proportion of actual positives correctly identified. The F1-score, a balanced measure, combines precision and recall, providing valuable insights into their interplay.

The Area Under the Receiver Operating Characteristic curve (AUROC) assesses a model’s ability to distinguish between negative and positive cases across a range of threshold values. In binary classification tasks, AUROC stands as a valuable assessment metric.

These metrics, including F1-score, precision, recall, and AUROC, play a pivotal role in the comprehensive evaluation of machine learning model performance, particularly in tasks like abnormality detection.

## V. EXPERIMENT

### A. The Training Process

In the experimental phase of our study, we utilized the TNMG subset data as the primary training dataset for our S4D-CfC models. Within this phase, comprehensive training and validation procedures were meticulously conducted. To effectively mitigate the class imbalance challenge inherent in multi-labeled datasets, we introduced a class weighting mechanism. Specifically, a class weight of 6 was thoughtfully assigned to the positive cases. This strategic adjustment was implemented to ensure a sufficient representation of positive-labeled data points within the training dataset, thereby rectifying the imbalance in the distribution of positive and negative labels.

The training of our models was conducted on a Tesla V100S-PCIE-32GB GPU. Throughout the training process, we employed a batch size of 32 to manage data processing efficiently. The training regimen comprised 300 epochs, and we maintained a learning rate of 0.003. To optimize the training process, we utilized the Adam optimizer in conjunction with the binary cross-entropy loss function. Additionally, a learning rate scheduler was incorporated into the training pipeline to enhance learning dynamics. This learning rate scheduler, known as the Cosine Annealing Learning Rate Scheduler, progressively decreases the learning rate over epochs using a cosine annealing pattern. This approach aids in model convergence by facilitating smoother adjustments to the learning rate, preventing abrupt changes that could hinder training stability and convergence.

### B. Assessing the Impact of Max-Min Scaling

Furthermore, we will investigate the influence of applying max-min scaling to the data on the model’s performance. This analysis will involve examining how the model’s predictive accuracy is affected when the ECG data is subjected to max-min scaling. By conducting experiments that encompass scaled and unscaled data, we aim to discern the consequences of this preprocessing step on the model’s effectiveness.

This investigation into the effects of max-min scaling on our model’s performance will provide valuable insights into the significance of data preprocessing techniques and their potential impact on model outcomes.

### C. Training and Validation Processes Within the Sample

For the assessment of model performance, a distinct dataset containing 20% of the TNMG subset was meticulously set aside for validation.

These selected data points were intentionally withheld from the model’s training phase to evaluate its effectiveness. This validation procedure was carried out separately for each model, enabling a direct and unbiased comparison of their individual performances.

### D. Radxa Zero Edge-AI Deployment

To validate the practicality of our wearable device model concept, we deployed the S4D-CfC model on the Radxa Zero. The Radxa Zero has an Amlogic 905Y2 processor featuring 64-bit ARM architecture and offers up to 4GB of 32-bit memory.

The pivotal on-chip fine-tuning of the experimental model took place on the Radxa Zero platform. Initially, we selected a model that had been pre-trained for 10 epochs on the TNMG dataset using a GPU, which served as the foundational model for fine-tuning. Given the limited computational resources available on the Radxa Zero, we implemented optimizations to ensure energy efficiency, thereby enabling the deployment of both the model and data on the device.

To facilitate this process, we employed a dataset comprising 1280 data points for fine-tuning, spanning a total of 50 epochs. Within this dataset, 20% of data was reserved for test and validation purposes. This fine-tuning procedure aimed to replicate the concept of model personalization based on new personal data inputs, allowing the model to adapt and enhance its performance in real-world scenarios.

### E. Evaluation on Unseen Data

We will rigorously evaluate the performance of our models using the CPSC dataset, meticulously chosen to mirror realworld scenarios. This evaluation entails comparing the models’ predictions against known values, employing a range of performance metrics. Through this comprehensive analysis, we will assess the strengths and limitations of the models, providing valuable insights that will inform decisions regarding their effectiveness. Additionally, these findings will serve as a foundation for identifying potential avenues for improving ECG analysis.

### F. Assessing Model Robustness

To gauge the robustness of our model, we will subject it to a battery of tests involving the deliberate removal of channels from the 12-lead ECG data. This evaluation aims to ascertain the model’s performance in the presence of missing inputs. The tests will be conducted in a progressive manner, systematically removing different numbers of channels, ranging from 1 to 6 leads. These experiments will gauge the model’s capacity to sustain accuracy in diverse scenarios.

We will employ a suite of performance metrics to measure its performance quantitatively. These metrics will enable us to evaluate and compare the model’s performance across various scenarios. The insights gained from these evaluations will be instrumental in identifying potential areas for model improvement and offer valuable guidance for future enhancements.

### G. Single Lead ECG Model Evaluation

As part of our comprehensive study, we specifically explored the model’s effectiveness in handling single lead ECG, focusing on Lead II. The model underwent specialized training using exclusive Lead II data from the dataset. We then evaluated its performance solely on single lead ECG, comparing it with the model trained on the entire 12-lead ECG. It’s important to note that the training process and model architecture are consistent with the 12-lead examination. The only difference lies in using Lead II ECG data for training and testing, providing a targeted understanding of the model’s capabilities in this specific context.

### H. Model Size Evaluation

In addition to our core analysis, we experimented to assess the model’s potential applicability in the tinyML domain. To achieve this, we varied the length of the ECG data in trials to minimize model latency. Additionally, we manipulated the model dimensions to explore the correlation between model performance and model size (number of parameters). This investigation aims to determine the model’s viability in tinyML scenarios, where resource constraints exist when the model runs on hardware.

## VI. RESULTS

In this section, we will offer a comprehensive analysis of the models’ performance. Additionally, our discussion will encompass an in-depth exploration of their generative capabilities and robustness. The results obtained from this model will be subjected to a comparative analysis against the performance of the ConvCfC model^17^ in the upcoming discussion section, facilitating a comprehensive discussion of their respective strengths and limitations.

### A. Performance Evaluation and Scaling Impact

The central thrust of this research revolves around the training of our proposed models, a process meticulously centered on the TNMG subset dataset, which has been extensively elucidated in prior sections of this paper. The TNMG subset dataset serves as the bedrock upon which the development and evaluation of our models rest. This training journey spanned 300 epochs, punctuated by diligent monitoring and meticulous metric recording. Critical performance metrics, including accuracy and loss, were consistently tracked for both the validation and training sets after the completion of each epoch.

In addition to the aforementioned, we introduced a data preprocessing step by applying max-min scaling to the raw data. This was undertaken to discern the potential impact of data scaling on the model’s performance. Notably, both the models’ performances on the original (raw) and scaled data were assessed using identical experimental settings. The outcomes of these assessments are thoughtfully illustrated in Figure 4, providing a comparative analysis of the model performances under the two distinct data conditions.

**FIG. 4.**
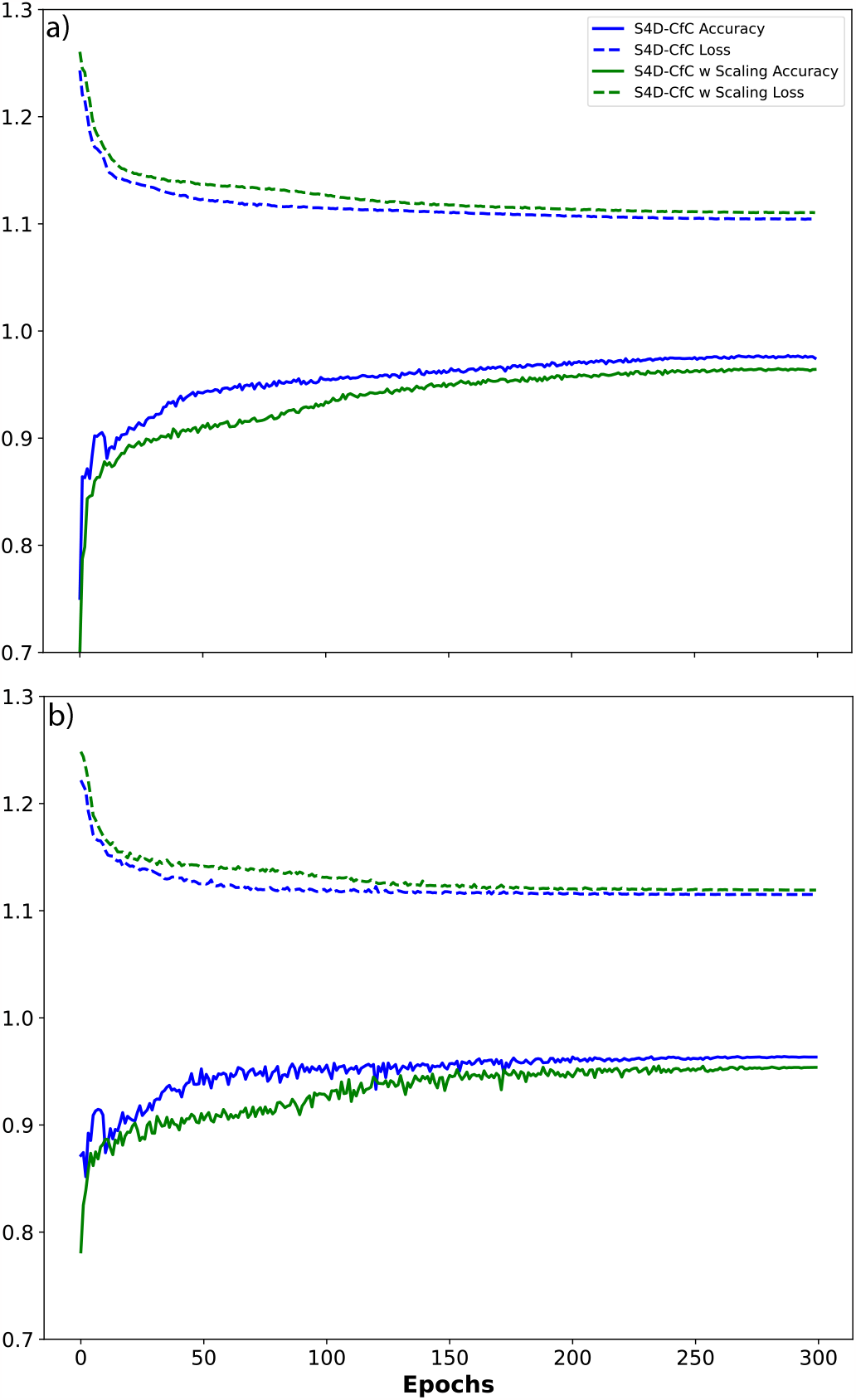
a) The figure provides a visual representation of the performance of models trained on both raw and scaled data throughout the training process. b) The figure presents a comprehensive view of the model’s performance by illustrating the trends in loss and accuracy metrics during the validation phase.

Figure 4(a) illustrates a consistent decrease in training loss as the training progresses, indicating the model’s improving performance over time. Concurrently, the training accuracy steadily rises, ultimately stabilizing and confirming the model’s effective learning from the training data. Notably, while both models, with and without data scaling, yield similar results, it’s worth noting that the model trained on raw data achieves a slightly higher accuracy compared to its scaled counterpart.

During the training phase, there is a consistent upward trend in validation accuracy, indicating an improvement in the model’s ability to generalize to unseen data. Moreover, the validation loss steadily decreases throughout the training period, reflecting the model’s evolving capability to make predictions that closely match the actual ground truth labels during validation. Figure 4(b) demonstrates that both models, whether trained on raw or scaled data, exhibit similar behavior. However, it’s noteworthy that the model trained on raw data consistently maintains a slight performance advantage over its scaled counterpart.

The model’s performance on raw and scaled data is succinctly presented in Table I. In both cases, the model demonstrates comparable performance, with an F1 score of 0.88 for the raw data model and 0.86 for the scaled data model. Additionally, the Area Under the Receiver Operating Characteristic (AUROC) values for raw data and scaled data are 0.98 and 0.97, respectively. These performance metrics corroborate the trends observed in Figure 4.

### B. On-Device Fine-Tuning

In this research endeavor, the S4D-CfC model was effectively deployed on the Radxa Zero, which is equipped with an Amlogic 905Y2 processor and boasts 4GB of memory. The fine-tuning procedure on the Radxa Zero was carried out utilizing a pre-trained model, which had initially undergone 10 epochs of training on a substantial dataset known as TNMG, with the computational assistance of a GPU. It is worth noting that the resultant saved model exhibited remarkable compactness, occupying a mere 242KB of storage space. Furthermore, the fine-tuning process demonstrated a noteworthy degree of memory efficiency, efficiently utilizing approximately 70% of the available memory resources.

The fine-tuning procedure entailed the utilization of a dataset comprising 1280 data points, spanned over 50 epochs, with subsequent validation conducted on 20% of the data points. The evaluation of the fine-tuned model revealed discernible improvements in performance metrics throughout the training process. Specifically, the average F1 score exhibited an increase from 0.48 to 0.56, and the Area Under the Receiver Operating Characteristic (AUROC) improved from 0.75 to 0.82. These findings underscore the efficacy of on-device fine-tuning in enhancing model performance. By conducting the fine-tuning process directly on the hardware, in this case, the Radxa Zero, tangible evidence of its positive impact on model performance is clearly demonstrated.

On-device fine-tuning serves as a testament to the adaptability and resource efficiency of the S4D-CfC model, highlighting its capability not only for initial training but also for subsequent fine-tuning within the resource-constrained computational environment of the Radxa Zero.

### C. Model Generalization

To thoroughly evaluate the adaptability of our models, we conducted predictive assessments on the CPSC dataset. These assessments were conducted using models that had been previously trained on the TNMG subset, as detailed in the data section. This evaluation serves as a rigorous test to assess how effectively the models can handle new and previously unseen data from the CPSC dataset, which inherently presents differences from the data on which they were trained. It is crucial to underscore that the CPSC dataset encompasses eight distinct types of abnormalities, with only four of them overlapping with those found in the TNMG dataset.

Through a meticulous analysis of our selected model’s performance on this CPSC dataset, we gain invaluable insights into its ability to transfer acquired knowledge effectively to novel and unfamiliar data scenarios. This analysis serves as a demanding benchmark to assess the model’s potential for generalization beyond its training data, enabling it to adapt to a broader range of clinical scenarios.

The performance of both models on unseen data has been meticulously summarized in Table I, vividly demonstrating their commendable performance. The model trained on raw data achieves an average F1 score of 0.76 and an AUROC of 0.93, while its counterpart trained on scaled data attains an F1 score of 0.78 and an AUROC of 0.93. These results underscore the robust generalization capabilities of both models. Intriguingly, it is worth noting that the model trained on scaled data exhibits slightly superior generalization performance compared to its raw data counterpart. This observation contradicts the findings from the evaluation phase, indicating nuanced variations in different facets of model performance.

**TABLE I.**
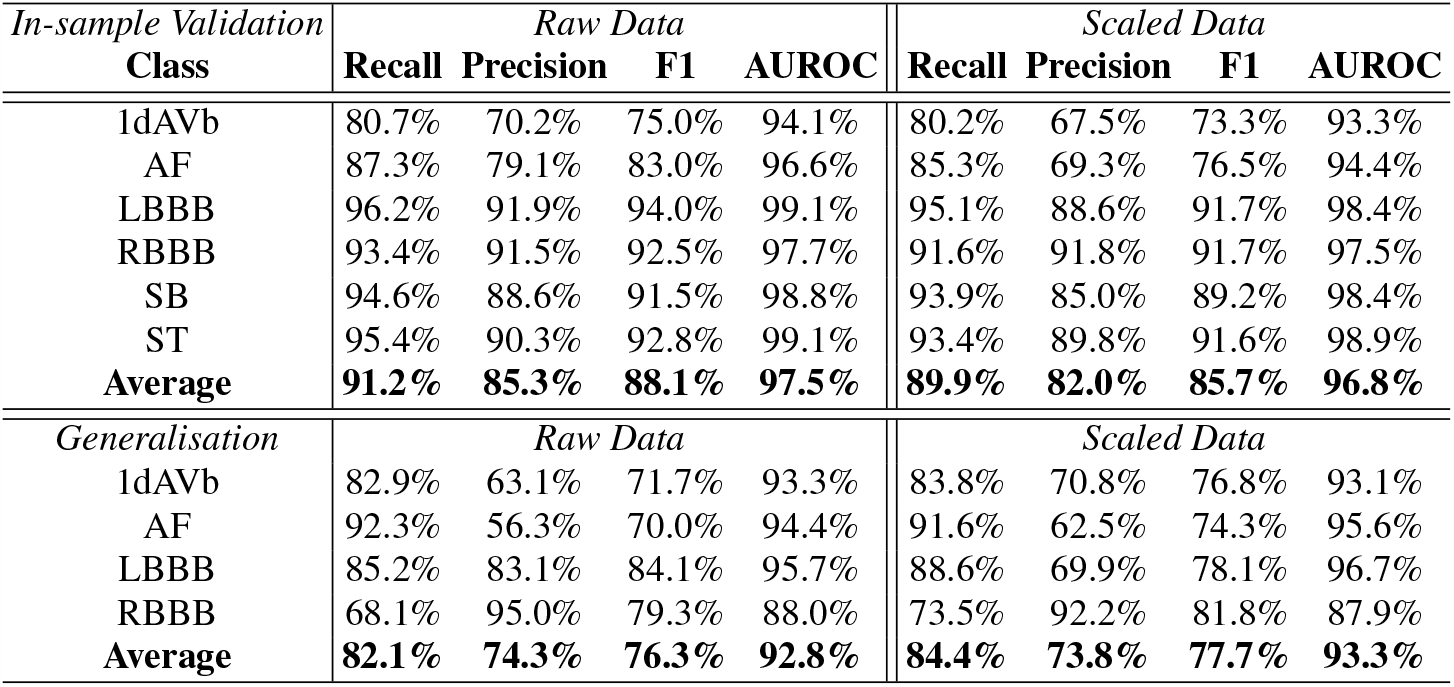
In-sample Validation and Generalisation Results for Model Trained on Raw and Scaled Data.

### D. Resilience of the Model

To comprehensively evaluate the model’s performance, we extended our assessments to incorporate the CPSC dataset, introducing deliberate variations. Specifically, we conducted additional evaluations by randomly removing specific channels from the 12-lead ECG data. This deliberate manipulation of the input data enabled us to thoroughly examine the model’s robustness and its capacity to effectively handle incomplete or missing input information. These experiments provide valuable insights into the model’s adaptability and resilience, especially in real-world scenarios where data integrity may encounter challenges.

In Figure 5, one can discern that as the quantity of omitted leads grows, the F1 performance metric of the models experiences a noticeable decline. It’s worth highlighting that the model trained on scaled data consistently outperforms its counterpart trained on raw data. This underscores a significant enhancement in model robustness achieved through data scaling, representing an intriguing and noteworthy discovery.

**FIG. 5.**
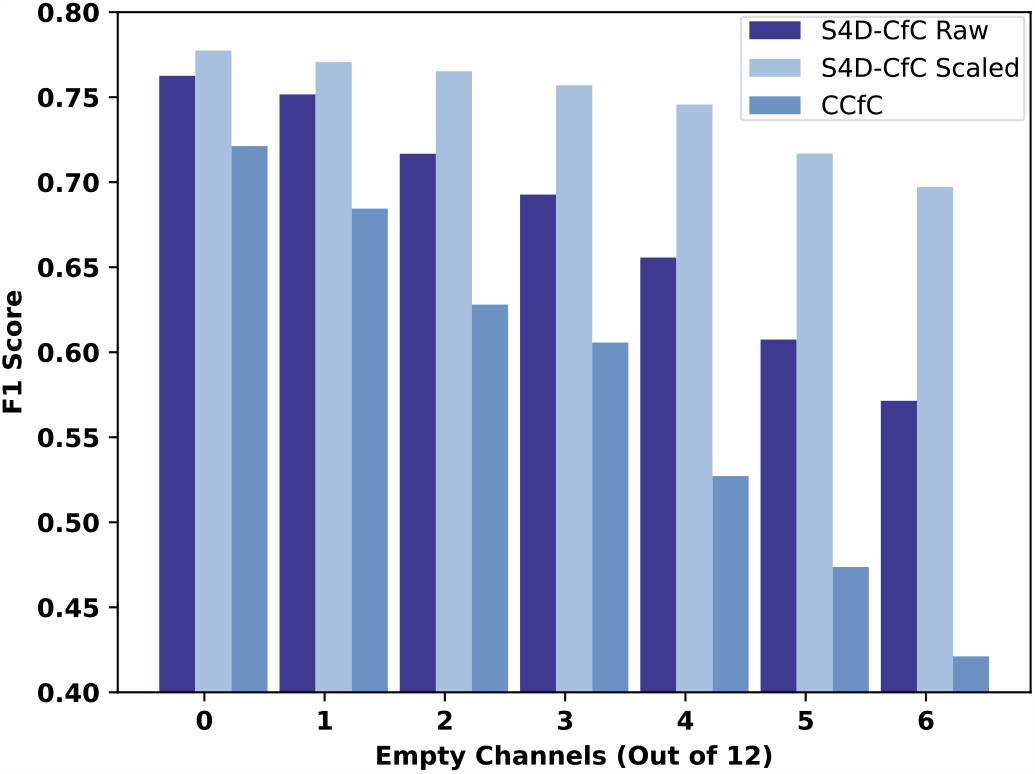
Evaluating model robustness via F1 metrics with varying degrees of missing channels in CPSC ECG data. The ConvCfC model^7^ serves as the baseline for assessment.

### E. Model Performance on Single-Lead ECG Data

As part of the study showcasing the model’s proficiency in processing single-lead ECG data, the model underwent exclusive training using both raw and scaled Lead II ECG data from the TNMG subset. The validation results of the model have been concisely presented in Table II.

The model consistently showcases satisfactory performance, emphasizing its proficiency in managing reduced-lead ECG data and signaling promising potential for future applications. Notably, the model trained on scaled single-lead ECG outperforms its counterpart trained on raw single-lead ECG, reflecting a slight improvement in performance.

**TABLE II.**
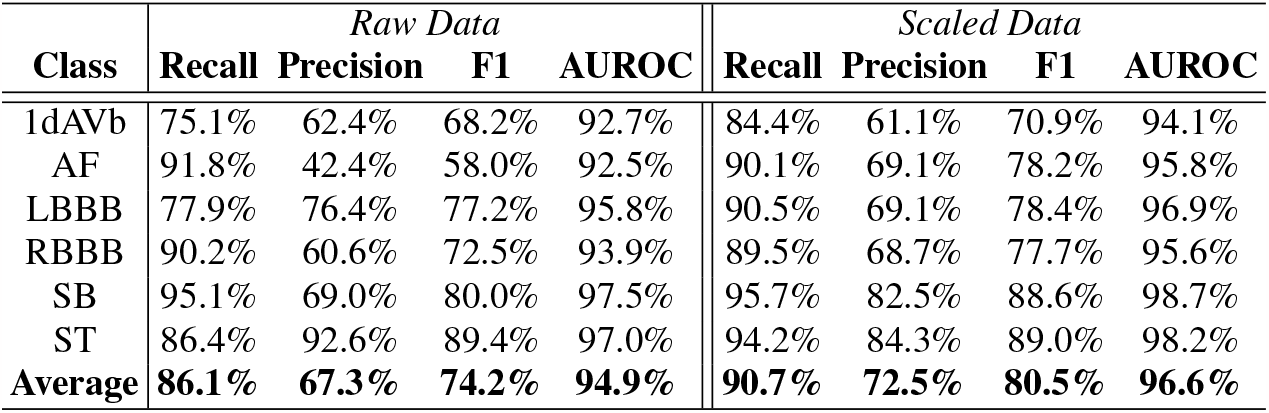
In-Sample Validation Results for Model Trained on Both Raw and Scaled Lead II ECG Data.

### F. Size Matters

In our study, we have discovered that the hyper-parameter known as “Model Dimension” (d_model) in the S4D layer plays a crucial role in both model performance and the number of parameters, which ultimately determines the model’s size. Given that model performance and efficient deployment on hardware are heavily dependent on model size (the number of parameters) and latency, we have also conducted investigations into the effects of reducing the input data length on model performance. The results of these experiments are summarized in Figure 6. The original data had a duration of 10 seconds, and by utilizing a shorter input length, we can effectively reduce latency, as the model is now processing data in less than the original 10-second time frame.

**FIG. 6.**
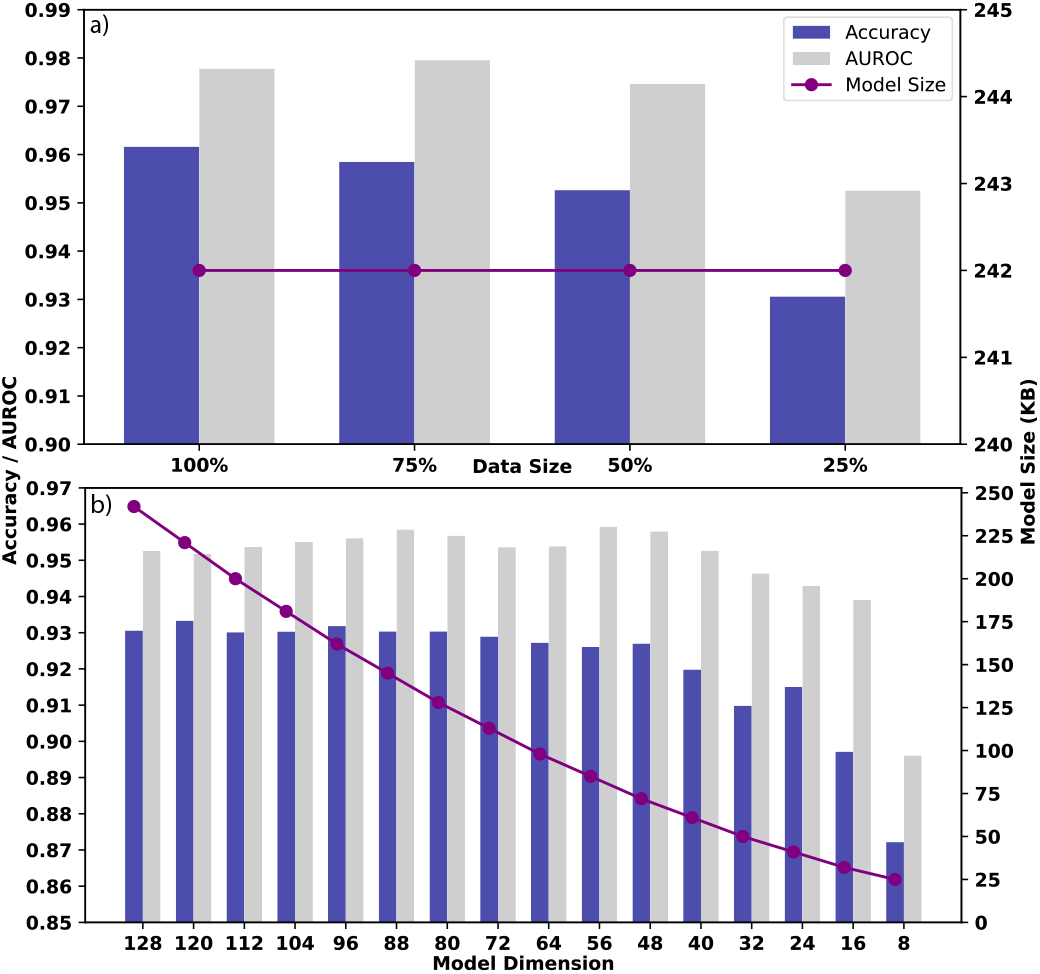
a) The chart illustrates the performance and model size changes resulting from training the model with only a fraction of the original 10-second input data time frame. b) The graph depicts the influence of d_model on both the model’s performance and its overall model size. These results were obtained by training the model with only 25% of the original 10-second data time frame.

It is evident that reducing the input data size has no noticeable impact on the model’s size, but it does affect its performance. Generally, the shorter the data length, the worse the performance, although the performance was not significantly affected until only 25% (2.5 seconds) of the data length was used. Despite a reduction in performance, it remains within an acceptable range, especially when considering the substantial 75% reduction in latency.

Conversely, the performance of the model exhibited a more pronounced sensitivity to d_model hyper-parameter. The model’s size is directly linked to d_model, and this correlation significantly influences the model’s performance. Initially, both the model’s performance and size increased with higher values of d_model. However, this trend eventually plateaus, with further increases in d_model having diminishing returns on performance. As the model size still scales proportionally with d_model, controlling the d_model effectively governs the model’s size.

Nevertheless, even with a mere 25 KB model size, the model still delivers acceptable performance when handling just 2.5 seconds of the original 10-second data length. This could make it an excellent candidate for implementation on extremely resource-constrained hardware, like microcontrollers, where both latency and model size (number of parameters) are crucial considerations.

## VII. DISCUSSION

In this section, we delve into the study’s results, emphasizing its findings regarding performance, generalization, robustness, and its capacity for on-chip learning. In this section, we will also employ the ConvLSTM2D CfC (ConvCfC) model as a baseline for comparison^7^. The ConvCfC model shares a similar structure to the model, with the difference being that the ConvLSTM2D layer in the S4D-CfC model is replaced by a layer of S4D. The performance of the ConvCfC model in both in-sample and generalization scenarios is presented in Table III.

**TABLE III.** Results from the ConvCfC Model^7^.

|  | Precision | Recall | F1 | AUROC |
| --- | --- | --- | --- | --- |
| Validation | 85.9% | 80.6% | 82.8% | 96.3% |
| Generalisation | 75.5% | 73.9% | 72.1% | 91.3% |

Regarding the validation performance of the models, both the S4D-CfC models, one trained on raw data and the other on scaled data, exhibited excellent results in terms of F1 scores and AUROC values. Notably, the model trained on raw data showed slightly superior performance compared to its scaled counterpart, particularly evident in the F1 Score and precision metrics, where the S4D-CfC model trained on raw data outperformed the scaled data-trained model. However, when comparing these results to those of the ConvCfC model, both S4D-CfC models have surpassed the ConvCfC model’s performance.

Table IV further juxtaposes the performance of the proposed S4D-CfC model with that of other works deploying the same dataset, underscoring the superior performance of the former.

**TABLE IV.** Evaluating Model Performance Using the TNMG Dataset.

| Ref | Model | F1 | AUROC |
| --- | --- | --- | --- |
| 10 | DL | – | 87% |
|  | FE | – | 86% |
|  | FE+DL | – | 86% |
| 11 | DNN | 0.20 | 91% |
| 12 | DNN | 0.88 | – |
| 13 | 1D-CNN | 0.80 | – |
| 7 | ConvCfC | 0.83 | 96% |
| **Proposed** | **S4D-CfC** | **0.88** | **98%** |
| **Proposed** | **S4D-CfC w/ Scaling** | **0.86** | **97%** |

However, the situation shifts when it comes to generalization on CPSC dataset. In this context, the S4D-CfC model trained on scaled data has slightly outperformed its counterpart trained on raw data. This behavior contrasts with the results we just discussed for in-sample testing, indicating a beneficial impact of data scaling when it comes to generalization. Furthermore, when we compare the generalization performance of the S4D-CfC models to that of the ConvCfC models, both S4D-CfC models have surpassed the performance of the ConvCfC model, once again underscoring the superiority of the proposed model.

During the robustness testing phase, some unexpected performance trends have emerged. While both S4D-CfC models consistently outperform the ConvCfC model, as illustrated in Figure 5, the performance of the S4D-CfC model trained on raw data and the one trained on scaled data has exhibited distinct behavior. Although the performance of both models declines as the number of empty channels used in inference increases, the model trained on scaled data has demonstrated significantly greater resilience and robustness when dealing with empty channels. This discovery holds significant implications for modelers, emphasizing the importance of considering data scaling strategies during model training to achieve better generalization and resilience.

The successful deployment of the S4D-CfC model on the Radxa Zero signifies a significant achievement in this research effort. This deployment underscores the model’s adaptability to resource-limited environments and its potential for practical applications. The fine-tuning process, which leveraged a pretrained model and a relatively small dataset, yielded impressive results in terms of compactness and memory efficiency. Notably, the fine-tuned model occupied just 242KB of flash memory and efficiently utilized approximately 70% of the available RAM resources, showcasing its efficiency.

Moreover, the observed enhancements in performance throughout the fine-tuning process are significant. The rise in the average F1 score from 0.48 to 0.56 and the improvement in the Area Under the Receiver Operating Characteristic (AUROC) from 0.75 to 0.82 underscore the effectiveness of on-device fine-tuning in elevating the model’s performance. This serves as a compelling proof of concept, demonstrating the model’s capacity for deployment on edge devices and subsequent personalized fine-tuning. This capability opens the door to a future where wearable medical devices can offer more personalized and tailored solutions, marking a promising direction in the evolution of healthcare technology.

## VIII. CONCLUSION

In conclusion, the outcomes of this study underscore the effectiveness and promise of the proposed models for identifying abnormalities. These models have exhibited strong performance, robust generalization capabilities, and the capacity to handle situations involving incomplete or missing input data. Additionally, the study has highlighted the notably beneficial impact of data scaling on both generalization and model robustness, particularly when dealing with missing data during inference. Furthermore, the research reaffirms the models’ effectiveness in hardware deployment, emphasizing their compact size and potential for on-device fine-tuning, a step closer to personalization in medical devices.

## Data Availability

This research paper uses the openly available CPSC dataset (http://2018.icbeb.org/Challenge.html) for analysis and experimentation. However, it is crucial to emphasize that the TNMG dataset, in contrast, is not open to the public. To access the TNMG dataset, one must secure permission from the data owner. While access may be granted upon approval, it is essential to highlight that the TNMG dataset is not readily accessible to the broader public.

## IX. ACKNOWLEDGEMENT

Zhaojing Huang expresses gratitude for the assistance and support the Australian Government’s Research Training Program (RTP) generously provided.

## X. CODE ACCESS

You can access the code via the link https://github.com/NeuroSyd/S4D-CfC. Kindly be aware that there could be specific terms, conditions, or usage restrictions applicable to the code.

## XII. CONFLICT OF INTEREST STATEMENT

The authors affirm that they have no conflicts of interest to disclose, including both financial and non-financial considerations.

## Notes

### Competing Interest Statement

The authors have declared no competing interest.

